# The effect of short-term and high-intensity exercise training in plasma lipidome profiles of people living with and without HIV

**DOI:** 10.1101/2025.06.02.25328634

**Authors:** Marcos Yukio Yoshinaga, Flávio Gomez Faria, Adriano de Britto Chaves-Filho, Sayuri Miyamoto, Tania Cristina Pithon Curi, Giselle Cristina Bueno, Bruno Ferrari Silva, Sidney Barnabe Peres, Solange Marta Franzói de Moraes

## Abstract

Both HIV infection and antiretroviral therapy contribute to dyslipidemia and abnormal body fat distribution in people living with HIV (PLWH). Exercise training is an effective intervention to protect against these metabolic changes. However, little is known about the mechanisms underlying the impact of exercise training on lipid metabolism in PLWH. To better understand lipid alterations, this study aimed to comparatively evaluate the effect of high-intensity-functional circuit training for 8 weeks on the plasma lipidome of PLWH (n=13) and HIV-negative subjects (control group; n=14). Anthropometric and biochemical parameters revealed lower levels of leptin, HDL-C, body fat %, and BMI combined with elevated Aspartate Transaminase (AST) and Homeostasis Model Assessment of β-cell function (HOMA_beta) in PLWH when compared to control subjects that persisted from baseline to post-exercise training. Nonetheless, contrasting levels of adiponectin, fasting insulin, HOMA_IR and Adipo_IR, together with phosphatidylcholine-containing lipids observed at baseline were equalized after training in PLWH. In control subjects, significant reductions in concentrations of triglycerides alongside phosphatidylinositol and glycosylated ceramides were observed post exercise training. By contrast, PWLH displayed a decrease in concentrations of free fatty acids, cholesteryl esters, and glycosylated ceramides together with increased levels of diglycerides, acylcarnitines, and free cholesterol after exercise training. In addition to specific lipidome alterations in each group, this study showed concomitant modulation of several glycerophospholipids and sphingolipids suggesting health-promoting effects of short-term high-intensity exercise training. Collectively, these modulated lipid species represent interesting targets for future lipidomic-based studies evaluating not only the effects of exercise training, but also the molecular mechanisms resulting in a healthier plasma lipidome profile.

## Introduction

Contemporary antiretroviral therapy (ART) has improved both survival and quality of life of people living with the human immunodeficiency virus (PLWH) (Palella et al., 2006; Feinstein et al., 2016). Nonetheless, dyslipidemia and lipodystrophy are highly prevalent in PLWH, significantly contributing to an increased risk of developing cardiovascular diseases (CVD) when compared to the general population (Shah et al., 2018; Feinstein et al., 2019; Jones et al., 2024). Lipid profile abnormalities have also been associated with the prevalence of other metabolic disorders such as non-alcoholic fatty liver diseases in PLWH, which may in turn increase the risk of cardiovascular events (Crum-Cianflone et al., 2009; Welzen et al., 2019; Krishnan et al., 2023). Dyslipidemia in PLWH can occur due to certain types of ART, particularly the first-generation protease inhibitors that may lead to metabolic dysfunctions (e.g. high LDL-C, low HDL-C and insulin resistance; Grinspoon and Carr, 2005 Kumar and Samaras, 2018; Koethe et al., 2020). Therefore, the management of dyslipidemia in PLWH requires close attention considering the drug-to-drug interaction between ART medications and lipid-lowering therapies (Edelman et al., 2013; Giguère et al., 2019).

Lifestyle interventions through diet and/or prescription of an exercise regime could represent an effective and complementary alternative to drug-based therapies in PLWH for prevention and treatment of CVD risk. Promising improvements in immune function, gut microbiota, cardiorespiratory fitness, blood lipids, among others, are reported in short-term interventions with aerobic exercise, resistance exercise, and their combination (O’Brien et al., 2016; Ceccarelli et al., 2020; Oliveira et al., 2020; Ozemek et al., 2020). There is limited evidence concerning the effects of exercise on blood lipid panels of PLWH (Jaggers and Hand, 2016; Quiles et al., 2020). In general, the existing data from various protocols demonstrate increased concentrations of HDL-C with or without decreased levels of cholesterol and triglycerides as the major impacts of exercise training.

Lipids play a central role in atherosclerosis, from formation and progression of atherosclerotic plaque in the protracted subclinical phase to its subsequent disruption at an advanced stage or after a cardiovascular event (Finn et al., 2010). By screening a large number of circulating lipids, plasma lipidome profiling by lipidomic analysis may provide insights into the molecular mechanisms underlying atherosclerosis (Ekroos et al., 2020; Diaz et al., 2021). Lipidomic screening of blood lipids has shown promising results detecting prominent differences in lipidome composition between PLWH and HIV-negative controls (Wong et al., 2014; Bowman et al., 2019; Chai et al., 2019; Zhao et al., 2019), which may be useful for risk stratification in chronic diseases. Interestingly, a recent study applied targeted lipidomics to evaluate the effects of a supervised and mixed endurance/resistance exercise on serum lipidome of PLHW and HIV-negative controls, indicating a very distinct response between these populations (Bowman et al., 2022).

Despite the significant advances in the field of lipidomics during recent years, reliable quantification and identification of the human lipidome are still major challenges for clinical translation (O’Donnell et al., 2020; Gao et al., 2020). In large-scale lipidomics studies, targeted methods have been preferred over untargeted approaches due to high sensitivity and throughput in quantification of a selected list of molecules. Conversely, by facilitating discovery of lipid species, untargeted methods can provide a more unbiased picture of plasma lipidome, which depending on population is quite variable in composition (Aristizabal-Henao et al., 2020). The latter when applied to population studies, however, is largely dependent on software performance, which is currently unable to identify 40 to 60% of ions detected by mass spectrometry (Tsugawa et al., 2017).

The present study is aimed at describing the effects of 8 weeks of high-intensity-functional circuit training on the plasma lipidome of PLWH (n=13) and HIV-negative subjects (control group; n=14). For this purpose a combined untargeted/targeted lipidomic approach was applied to evaluate plasma lipidome profiles of these groups of subjects at baseline and after 8 weeks of exercise training.

## Methods

### Study design and participants

Sedentary adult men and women seronegative (HIV-) or diagnosed with HIV (HIV+) were recruited from the Specialized Care Service for STD/AIDS (Serviço de Assistência Especializada in Maringá, Paraná, Brazil). Participants willing to join the study signed the Free and Informed Consent Form and responded to an anamnesis form. The experimental study procedures were approved by the Institutional Research Ethics Committee under the registration n^o.^ 3211855 (Universidade Estadual de Maringá). The experimental trial followed the recommendations outlined by the CONSORT statement for non-pharmacological interventions (Boutron et al., 2008). The clinical trial was registered in the Brazilian Registry of Clinical Trials (UTN: U1111-1231-1846).

The inclusion criteria for PLWH were positive serology for the HIV virus and ART use for at least six months, lack of motor impairment or comorbidity to perform tests and training, lack of regular physical activity in the last six months, over 18 years of age and medical certificate for cardiometabolic capabilities to perform physical exercise. The exclusion criteria for both PLWH and HIV-negative controls were diarrhea, nausea, vomiting or poor oral food intake, systemic infection in the last 30 days, pregnancy and at least 85% of participation during training sessions.

The sample size was calculated using the G*Power software (Düsseldorf, Germany), considering the power of the test (1-β = 80%). From this estimation, the ideal size of the study was established at 34 participants. The present two-arm, parallel, non-randomized clinical trial evaluated the effect of an 8-week exercise training on plasma hormonal and biochemical responses in PLWH relative to HIV-negative subjects (Silva et al., 2022). From a total of 37 subjects submitted to eight weeks of exercise training program, the present study examined a subset of 27 participants with the focus on plasma lipidomic analysis. The experimental trial followed the recommendations outlined by the CONSORT statements for non-pharmacological interventions (Boutron et al., 2008), and professionals involved in the selection of the groups were not included in the evaluations and interventions, reducing the risk of bias. This clinical trial was registered in the Brazilian Registry of Clinical Trials (UTN: U1111-1231-1846).

### Functional exercise training program

To carry out the training, the following materials were used: mats, steps, TRX, flexion support, Swiss ball. The activities were presented in order to develop and stabilize the muscles of the central region of the body (abdominal, lumbar, pelvis and hip), improving strength and muscle power. In addition, exercises involving motor skills such as balance, coordination, gait, agility and proprioception were used.

The training program lasted eight weeks, distributed in three weekly sessions with a total duration of 5 minutes of warm-up, 30 minutes of the exercise execution, composed of ten exercise stations (jumping-up jumping jacks, push-ups, squats, lunges, hip raise, burpee, plank, sit-up, crate jump, bilateral back row). Two rounds were held in a circuit format; the execution time and interval between exercises were controlled in the proportion of 2:1 (effort: pause), being 1 minute of stimulus and 30 seconds of rest, followed by 5 minutes of stretching and cooling down.

### Anthropometric measurements, blood plasma sampling and biochemical analyses

The multifrequency, tetrapolar electrical bioimpedance technique (Inbody®, model R20; InBody Co., Seoul, South Korea) was used to measure body composition. The evaluations were carried out in the morning after fasting for at least 12 hours. Thereafter, approximately 8 mL of blood were drawn and distributed into tubes containing EDTA or heparin, which were centrifuged at 5,000 *g* for a period of 10 minutes. Aliquots of plasma were stored in 1.5 mL tubes and stored at −80° C until further analysis.

Plasma samples were used to determine the concentrations of glucose, glutamate-pyruvate transaminase (GTP), oxaloacetic transaminase (GOT), gamma glutamyl transferase (GGT), total cholesterol and triglycerides, very low-density (VLDL), low-density (LDL) and high-density (HDL) lipoproteins were determined by colorimetric methods using kits from Gold Analyze (Belo Horizonte, MG, Brazil). The following hormones were also determined: insulin, leptin and adiponectin. The concentrations of these hormones were measured by ELISA technique followed by analysis on a FlexStation3 plate reader (Molecular devices, USA) using a commercial kit (Invitrogen®, USA) and according to the manufacturer’s recommendations.

Insulin resistance was assessed via homeostasis model assessment of insulin resistance (HOMA_IR) using the formula: HOMA_IR = Fasting glucose (mg/dL) × fasting insulin (μU/mL) / 405 (Matthews et al., 1985). Values between 2.0–2.5 for HOMA_IR indicate healthy individuals and higher than 2.5 insulin resistance. The beta-cells function was evaluated using HOMA_BETA via formula: HOMA_BETA = 360 × (fasting insulin [μU/mL] / fasting glucose [mg/dL]) – 63. Reference values of HOMA_BETA for healthy individuals are between 100 and 150 with those higher than 150 indicating hyperinsulinemia (Matthews et al., 1985). Insulin sensitivity was also assessed via quantitative insulin sensitivity check index (QUICKI) using the formula: QUICKI = 1 / (log fasting insulin [μU/mL] + log fasting glucose [mg/dL]). Results from QUICKI of 0.33–0.36 are observed in healthy individuals, while values below 0.33 indicate insulin resistance (Katz et al., 2000). Finally, the insulin resistance in adipocytes was calculated via Adipo_IR index by the formula: Adipo_IR = free fatty acid (mmol/L) × fasting insulin (μU/mL). Adipo_IR values below 10 suggest healthy individuals, whereas those higher than 10 indicate high likelihood of metabolic syndrome (Gastaldelli et al., 2009).

### Lipidomic analysis

Plasma samples (50 µL) were extracted according to Matyash et al. (2008) with minor modifications and analyzed by a lipidomic analysis protocol developed by our group (Yoshinaga et al., 2021). A mix of internal standards (100 µL; **Table SX**) was spiked to the samples and afterwards 200 µL of cold methanol was added and this mixture was vortexed for 10 seconds. Subsequently, 1 mL of methyl tert-butyl ether (MTBE) was added and the mixture vortexed for 30 seconds and stirred at 1000 rpm for 1h at 20°C. In the following step, 300 µL of water was added to the tubes, which were kept in an ice bath for 10 min, and samples were vortexed (30 seconds) and centrifuged (10,000 g for 10 min at 4°C). After centrifugation, the supernatant containing the total lipid extracts (TLE) was collected and dried in a microtube under N_2_ gas. The TLE were resuspended in 100 µL of isopropanol and centrifuged (1500 g for 3 min at 4°C). Aliquots of 5 µL from all samples were collected in a vial and used as quality control sample, which was injected in the mass spectrometer at the beginning and after 9 experimental samples.

The TLE were injected into and lipids separation obtained by ultra-high pressure liquid chromatography (Nexera UHPLC, Shimadzu, Kyoto, Japan) equipped with a CORTECS® (UPLC® C18 column, 1.6 µm, 2.1 mm i.d. × 100 mm). Operating in a flow rate of 0.2 mL/min at 35 °C, the mobile phases consisted of solvent A (water and acetonitrile [60:40]) and solvent B (isopropanol, acetonitrile and water [88:10:2]). The gradient elution process was initiated at 40 to 100% of solvent B over the first 10 min. The samples were kept at 100% of solvent B from 10 to 12 min, and decreased to 40% of solvent B from 12 to 13 min, which was hold for 20 min. The UHPLC was coupled to a triple quadrupole time-of-flight mass spectrometry (Triple TOF® 6600, Sciex, Concord, US), with the electrospray ionization operating at −4.5kV and 5.5kV, for negative and positive ionization modes respectively, and cone voltage set at −/+80V. The curtain gas was set at 25psi, nebulizer and heater gases to 45 psi and interface heat a 450 °C. The scan range was set to 200–2000 Da.

The Information Dependent Acquisition (IDA) data were analyzed in PeakView software (Sciex, Concord, US). The identity of lipids was based on m/z, retention time, specific fragments and/or neutral losses in MS/MS (Han, 2016; Chaves-Filho et al., 2019). The identification of lipids in the untargeted approach aimed at 70 to 85% coverage of the most abundant ions in IDA. The annotated lipid identities and retention times were then used in the targeted approach, which consisted of peak areas of the precursor ions (±5 mDa) calculated using the MultiQuant software (Sciex, Concord, US). The peak areas of compounds of interest were then normalized by the peak area of the corresponding internal standard. The final concentration was given as nM lipid per µL of plasma (**Table SX**). The nomenclature of the fatty acyl chains of lipid subclasses are given as X:Y (with X representing the number of carbons and Y the number of double bonds). For sphingolipids, the sphingoid base is expressed as dX:Y and the n-acyl chains as X:Y. It is important to mention that the precise position of the fatty acyl chains in the glycerol backbone of glycerolipids and glycerophospholipids or the sugar moieties of hexosyl ceramides could not be exactly determined using our methods.

### Statistical analysis

The lipidomic analysis results were uploaded into MetaboAnalyst (https://www.metaboanalyst.ca/) (Chong et al., 2018). Prior to statistics, the coefficient of variation (CV) was calculated from quality control samples measured throughout the analytical experiments for each lipid reported in this study. Lipid species displaying a CV higher than 20% were excluded from statistics. For normal distribution, the data were log10 transformed prior to multivariate and univariate analyses. Multivariate analyses were performed by principal component analysis (PCA) and partial least square discriminant analysis (PLSDA). Both unpaired (to test baseline and post exercise differences between PLWH and controls) and paired (to test the effects of exercise training: post versus baseline within PLWH and controls) univariate analyses were conducted by t-test (p<0.05 for significance and false discovery rate applied). Univariate analyses of the anthropometric and biochemical data were performed using the same approach.

## Results

Lipidomic analysis was performed in a small subset of samples from a non-randomized trial, from which anthropometric and biochemical parameters have been reported elsewhere (**Silva et al., 2021; Melo et al. 2019; Guariglia et al. 2018**). To compare the lipidomic data with these clinical measurements, the major differences between PLWH and control subjects used in this study are described for baseline and post exercise training. At baseline conditions, PLWH presented significantly lower body fat %, BMI, HDL-C and leptin together with higher AST and HOMA_beta as compared to control subjects, and these parameters were maintained significantly different after exercise training (**Table 1**). In contrast, higher levels of adiponectin, insulin, HOMA_IR and Adipo_IR observed in PLWH relative to control subjects at baseline were no longer significant post exercise (**Table 1**). Nonetheless, in addition to the differences in several parameters that were maintained from baseline to post exercise training, levels of total cholesterol were reduced in PLWH as compared to control subjects after exercise training (**Table 1**).

**Table 1.** Anthropometric and biochemical parameters expressed as average ± standard deviation at baseline and post exercise training in PLWH and control subjects.

|  | Control (n=13) |  | PLWH (n=14) |  |
| --- | --- | --- | --- | --- |
| Age (years) | 37.76±14.1 |  | 40.7±16.0 |  |
| Female (%) | 77 |  | 57.2 |  |
| Anthropometric measurement |  |  |  |  |
|  | Baseline | Post | Baseline | Post |
| Weight (kg) | 82.9±23.5 | 82.7±22.3 | 69.5±26.1 | 69.2±25.3 |
| BMI | 29.8±6.6 | 29.9±6.4 | 24.7±8.1& | 24.6±7.9\$ |
| Eutrophic (%) | 30.8 | 30.8 | 46.1 | 46.1 |
| Overweight (%) | 23.1 | 23.1 | 46.1 | 38.5 |
| Obese (%) | 46.1 | 46.1 | 7.8 | 15.4 |
| Muscle (kg) | 29.2±6.3 | 28.9±6.2 | 27.7±9.8 | 27.8±9.7 |
| Fat (%) | 34.7±10.4 | 35.3±10.6 | 27.0±11.1& | 26.2±10.4\$ |
| Glucose and insulin homeostasis |  |  |  |  |
| Glucose (mg/dl) | 95.0±12.2 | 94.2±26.9 | 88.9±11.0 | 89.8±7.9 |
| HbA1 (%) | 5.2±0.4 | 5.3±0.5* | 5.30±0.23 | 5.2±0.3 |
| Insulin (μU/mL) | 10.6±5.3 | 9.5±3.5 | 27.4±21.5& | 9.3±4.8* |
| HOMA_IR | 2.5±1.3 | 2.2±0.7 | 6.46±5.44& | 2.0±0.9* |
| HOMA_beta | 170.5±159.9 | 145.5±125.5 | 372.3±172.2& | 352.3±239.7\$ |
| QUICKY | 0.34±0.02 | 0.34±0.02 | 0.32±0.04 | 0.35±0.02* |
| Adipo_IR | 11.0±7.5 | 10.0±5.6 | 27.2±24.7& | 11.4±10.1 |
| Biochemical measurements |  |  |  |  |
| Leptin (ng/mL) | 39.7±28.8 | 26.1±19.9* | 11.7±10.3& | 16.7±17.7\$ |
| Adiponectin (mg/mL) | 3.0±0.8 | 2.5±1.3* | 4.6±2.3& | 2.6±1.2* |
| AST (U/L) | 25.8±8.4 | 24.9±7.3 | 32.4±7.5& | 30.4±5.0\$ |
| ALT (U/L) | 28.1±16.4 | 26.2±20.4 | 42.2±23.9 | 31.7±11.8* |
| GGT (U/L) | 23.1±11.1 | 28.9±24.9 | 27.0±9.8 | 24.9±10.7 |
| AST/ALT | 1.04±0.4 | 1.19±0.5 | 0.93±0.4 | 1.07±0.4 |
| Adipo/Lept | 0.23±0.4 | 0.23±0.4 | 1.26±3.2& | 0.66±1.2\$ |
| Lipid panel |  |  |  |  |
| T_CHO (mg/dL) | 199.3±38.4 | 190.7±35.2 | 177.6±43.7 | 161.1±34.5*\$ |
| T_TG (mg/dL) | 151.5±77.7 | 114.3±61.6* | 119.1±61.4 | 136.4±84.1 |
| VLDL (mg/dL) | 30.3±15.5 | 22.9±12.3* | 23.8±12.3 | 27.3±16.8 |
| LDL-C (mg/dL) | 117.8±31.5 | 110.6±28.1 | 113.6±36.3 | 97.6±29.7* |
| HDL-C (mg/dL) | 53.0±11.1 | 54.1±12.1 | 41.1±9.9& | 39.9±8.6\$ |

\*Baseline vs post exercise; &Baseline (PLWH vs Control); \$Post exercise (PLWH vs Control); # The results are expressed in mean ± standard deviation.

This study identified and quantified a total of 418 individual plasma lipid species distributed into six major lipid classes: glycerolipids, glycerophospholipids, sphingolipids, fatty acyls, sterols and prenols (**Figure 1**). As commonly reported in untargeted lipidomic studies, triglycerides (TG) were the most diverse subclass with 134 lipid species (**Table SX**). The abundant TG species were followed by phosphatidylcholine (PC; 54 sp), sphingomyelin (SM; 34 sp), unsubstituted ceramides and free fatty acids (Cer and FFA, respectively; 24 sp each), and plasmenyl-PC and -phosphatidylethanolamine (pPC and pPE, respectively; 20 sp each) (**Figure 1**). Other subclasses composed of more than 10 lipid species included cholesteryl esters (CE; 19 sp), PE (16 sp), acylcarnitines (AC; 14 sp), phosphatidylinositol (PI; 13 sp) and plasmanyl-PC (oPC; 11sp). Lastly, lipid subclasses with relatively fewer molecular species comprised mono- and di-hexosyl ceramides (1H- and 2H-Cer with 9 and 2 sp, respectively), plasmanyl-PE (oPE; 4 sp), lyso-PC (LPC; 7 sp), lyso-PE (LPE; 4 sp), diacylglycerides (DG; 5 sp) and phytosterol esters (PhE; 2 sp), as well as single species of free cholesterol (FC) and coenzyme Q-10 (Q10).

**Figure 1.**
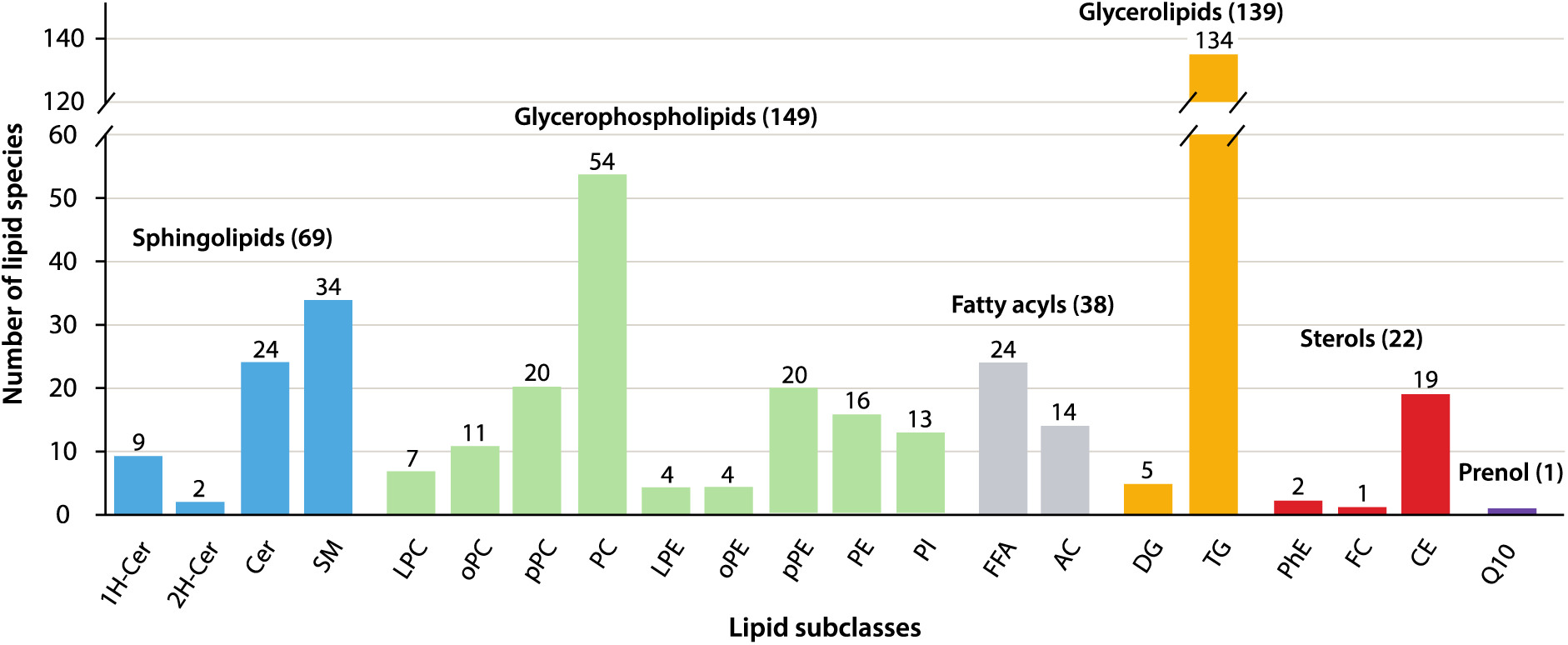
Plasma lipidome diversity displaying the total number of molecular species within each lipid class and subclass. Please see the main text for the description of lipid subclasses abbreviations.

**Table SX. Molar concentrations of all plasma lipid species (nM/L of plasma). Please see the main text for the description of lipid subclasses abbreviations.**

To examine the differences in plasma lipidome compositions of PLWH and control subjects, unpaired t-test coupled to fold-change analysis was performed at baseline and after training (**Figure 2**). Of note, with only a few exceptions, most plasma lipids exhibited <2-fold change. At baseline, 57 plasma lipid species were found in reduced concentrations in PLWH relative to control subjects. These altered lipids were mainly represented by pPC (6 sp), PC (15 sp) and SM (27 sp), comprising respectively 30, 28 and 79% of all species within these subclasses monitored in this study (**Figure 2a**). In comparison, after 8 weeks of high-intensity training, a relatively higher number of 77 plasma lipid species were found in altered concentrations between groups (**Figure 2b**). A major reduction was observed in 12 pPE species (representing 60% of all pPE species described in this study), with one of them exhibiting a >4-fold change between PLWH and control subjects. Interestingly, most of the down-regulated lipids in PLWH as compared to control subjects at baseline (particularly pPC, PC and SM) were normalized after exercise training, with only a few exceptions (**Figure S1**). Plasma lipid species displaying increased concentrations in PLWH relative to control subjects after exercise training represented 70% of all altered species (**Figure 2b**), and were represented by TG (41 sp), PI (5 sp) and DG (2 sp). Of note, one out of four PC species displayed >3-fold higher concentration in PLWH as compared to control subjects post exercise. Clearly, however, these results reflected the effects of exercise training in plasma lipidome of control subjects (e.g. decreased concentrations of TG, PI and PC) and PLWH (e.g. decreased concentrations of CE and increased levels of DG) relative to baseline, which will be described next.

**Figure 2.**
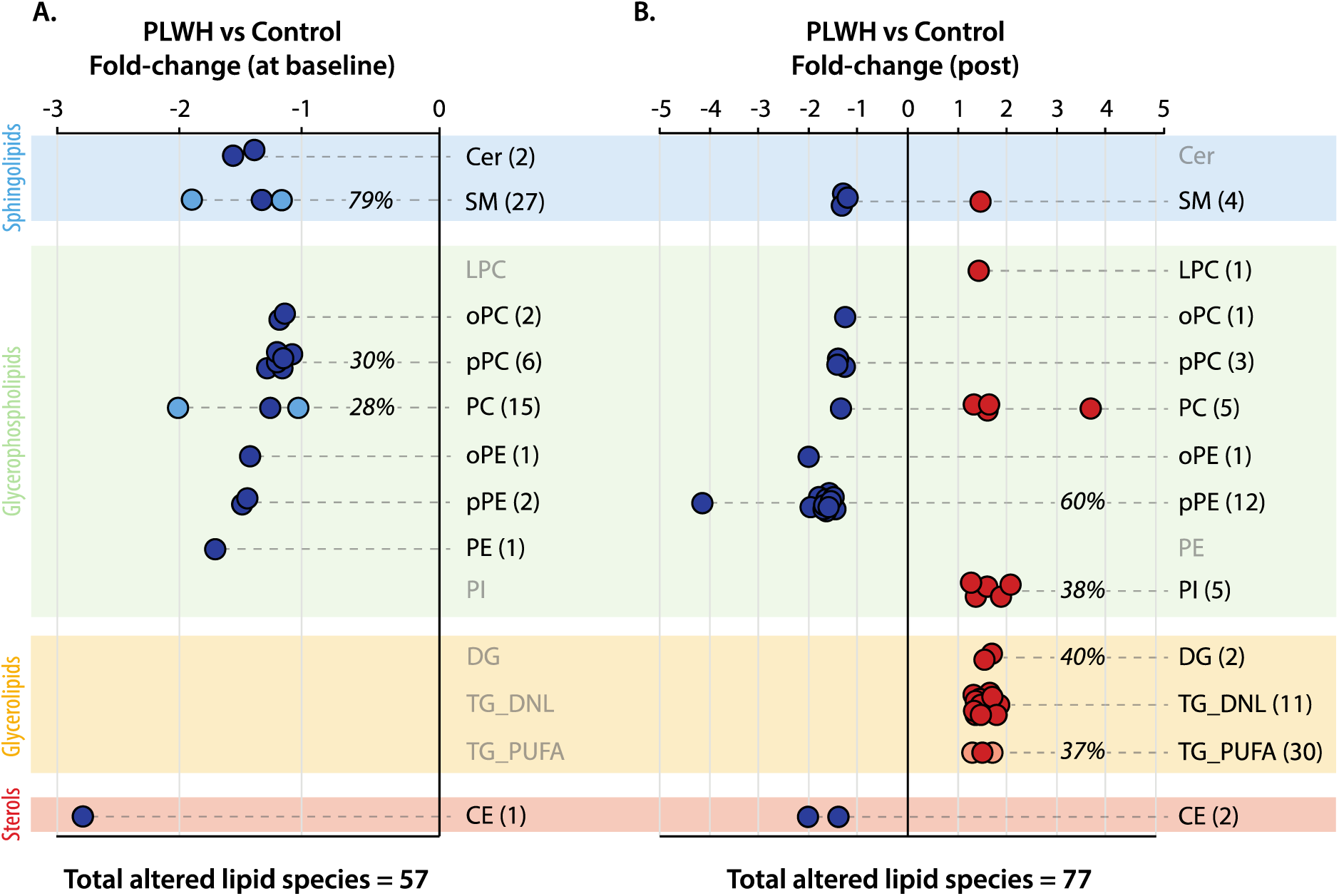
Fold-change values of significantly altered plasma lipid species differentiating PLWH and control subjects at baseline (a) and post exercise training (b). Background colors indicate distinct classes of lipids and are described accordingly (left corner). An unpaired t-test with p<0.05 for significance was applied for these comparisons. Please see the main text for abbreviations.

**Figure S1.**
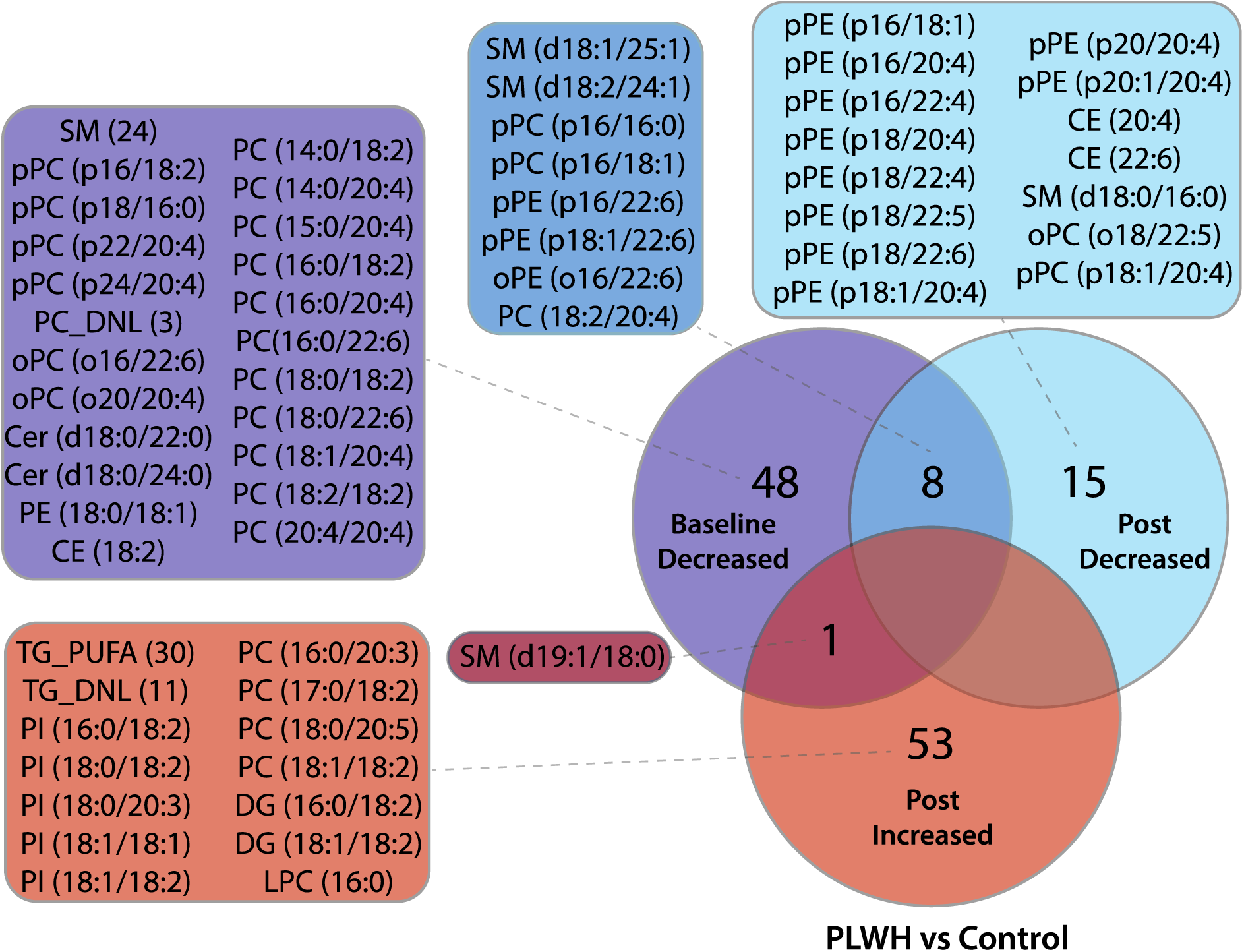
Venn diagram of the comparisons PLWH versus control subjects at baseline and post exercise (as illustrated in Figure 2). Please see the main text for abbreviations.

To examine the effects of exercise training in PLWH and control subjects both multivariate and univariate analyses were performed with paired data. Unsupervised and supervised multivariate analyses were performed by principal component analysis (PCA) and partial least square discriminant analysis (PLSDA), respectively. Both PCA and PLSDA revealed a moderate segregation of groups in score plots (**Figure S2a and S2c**). With a total of 64.3% and 59.2% explanation of data variability based on the two principal components of the PC and PLSDA, respectively, a consistent spatial segregation of exercise training versus baseline conditions was achieved in score plots (**Figure S2a and S2c**). The loadings plots showed that concentrations of glycerolipids, particularly TG, were dominant features regulating negative and positive values of component 1, respectively for PCA and PLSDA (**Figure S2b and S2d**). Positive values in component 2 of the PCA and PLSDA were determined by a handful of glycerophospholipids and 2H-Cer (d18:1/24:1), whereas negative values driven by glycerolipids concentrations (**Figure S2b and S2d**). It is worth mentioning that exercise training promoted a clear segregation of groups in PCA and PLSDA, particularly for control subjects (**Figure S2a and S2c**). Of note, suggesting a more pronounced reduction in plasma TG concentrations, most of the samples from the control group post exercise training scored positive and negative for component 1 of PCA and PLSDA, respectively, contrasting to samples from the PLWH group.

**Figure S2.**
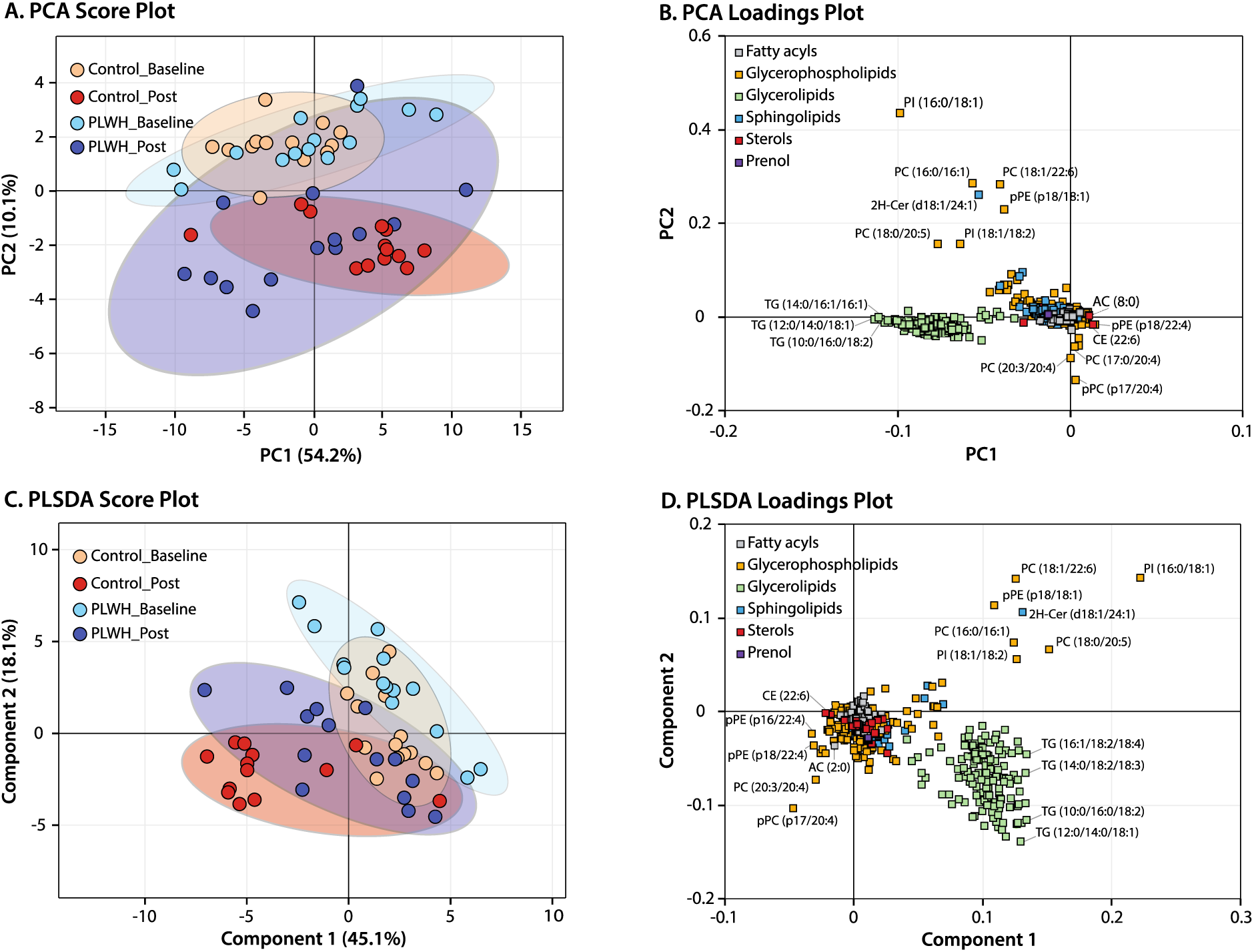
Results from multivariate analyses by principal component analysis (PCA) and partial least square discriminant analysis (PLSDA) showing score (A,C) and loadings (B,D) plots. Please see the main text for the description of lipid subclasses abbreviations.

Exercise training resulted in distinct lipidome profile alterations in PLWH and control subjects according to paired univariate analysis (**Figure 3**). In control subjects, exercise training led to significantly altered concentrations of 171 lipid species, most of them displaying reduced concentrations (164 sp) and 80% of them with >2-fold change as compared to baseline (**Figure 3a**). The data highlight that several TG and PI (representing 92 and 77% of all monitored species, respectively) along with 1H- and 2H-Cer (67 and 100% of total species, respectively) displayed reduced concentrations relative to baseline. Other modulated lipid subclasses with relatively high number of species included PC (9 sp) and SM (10 sp), with most of them displaying reduced concentrations in post exercise as compared to baseline. Of note, almost all PC species displayed >2-fold change, with three of them reaching 20-fold reduction post exercise (**Figure 3a**). In addition to PC, relatively high negative fold-change values were observed for two PI (9- and 59-fold) and single species of pPE (17-fold) and 2H-Cer (19-fold).

**Figure 3.**
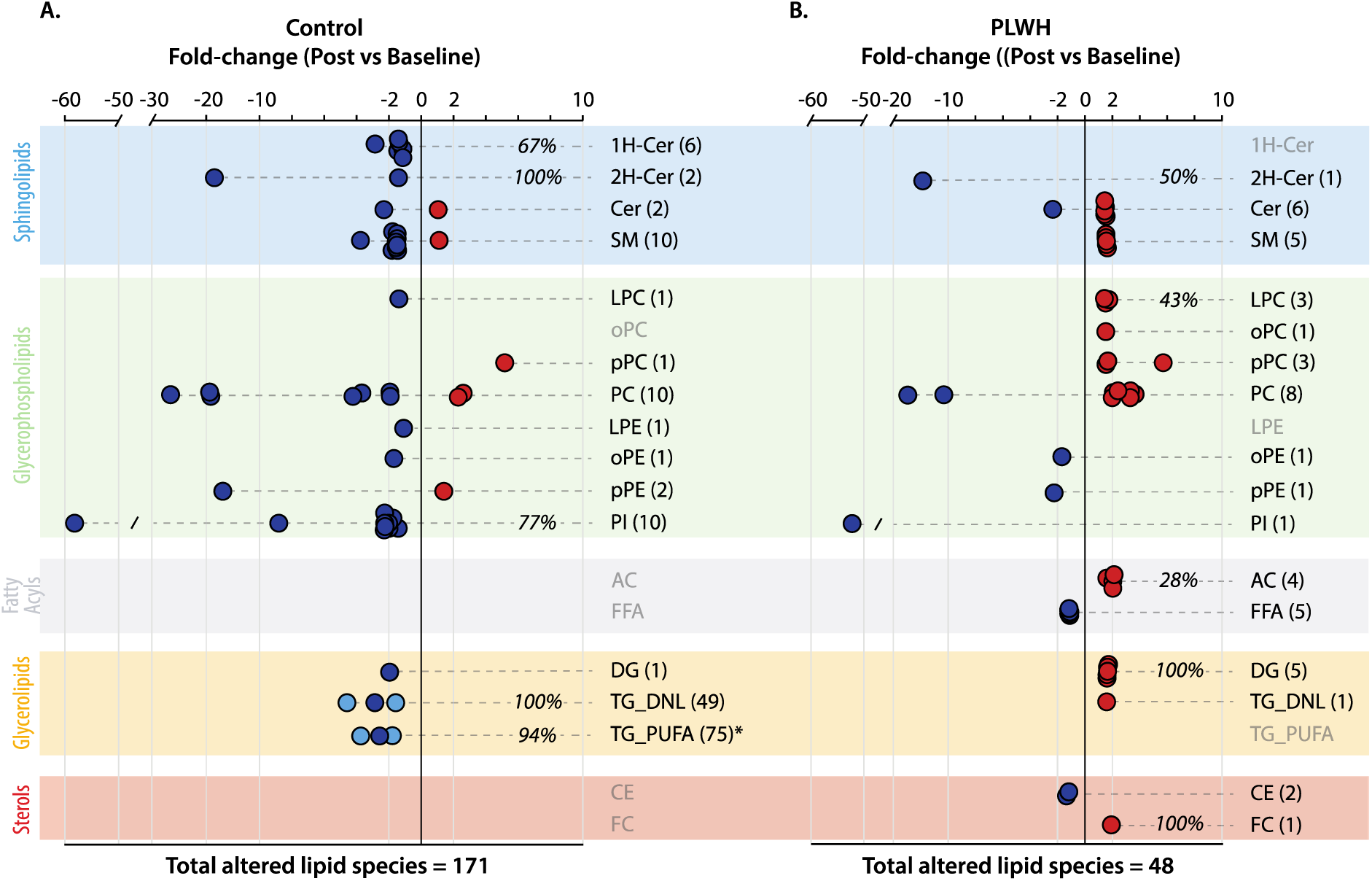
Fold-change values of significantly altered plasma lipid species differentiating post exercise and baseline in control subjects (a) and PLWH (b). Background colors indicate distinct classes of lipids and are described accordingly (left corner). A paired t-test with p<0.05 for significance was applied for these comparisons. Please see the main text for the description of lipid subclasses abbreviations.

As compared to control subjects, the modulation of plasma lipidome profiles by exercise training in PLWH was much less extensive, 47 species in total with only 20% of them displaying >2-fold change (**Figure 3b**). Unlike the alterations observed in control subjects, increased concentrations of plasma lipids prevailed after exercise training (33 species in total), including FC, DG, LPC and AC representing respectively 100, 100, 43 and 28% of all species monitored in this study. In addition, several plasma species of PC and SM in PLWH that were found in lower concentrations relative to control subjects at baseline (**Figure 2a and 2b**) displayed increased levels after exercise training (**Figure 3b**). Despite exhibiting a low number of species with significant reductions, the concentrations of 5 FFA (averaging 1.2-fold) and 2 CE (with 1.2- and 1.3-fold) were decreased after exercise training. In summary, a Venn diagram revealed very few overlaps in the modulation of plasma lipidome by exercise training in PLWH and control subjects, suggesting a distinct scenario with most of the changes linked to increased and decreased concentrations of unique lipid species, respectively (**Figure S4**). Interestingly, however, an overlap of four species with >10-fold reduction appeared as strong indicators of exercise training relative to baseline for both PLWH and control subjects, respectively: 11- and 27-fold for PC (16:0/16:1), 17- and 19-fold for PC (18:1/22:6), 51- and 59-fold for PI (16:0/18:1) and 14- and 19-fold for 2H-Cer (d18:1/24:1).

**Figure S3.**
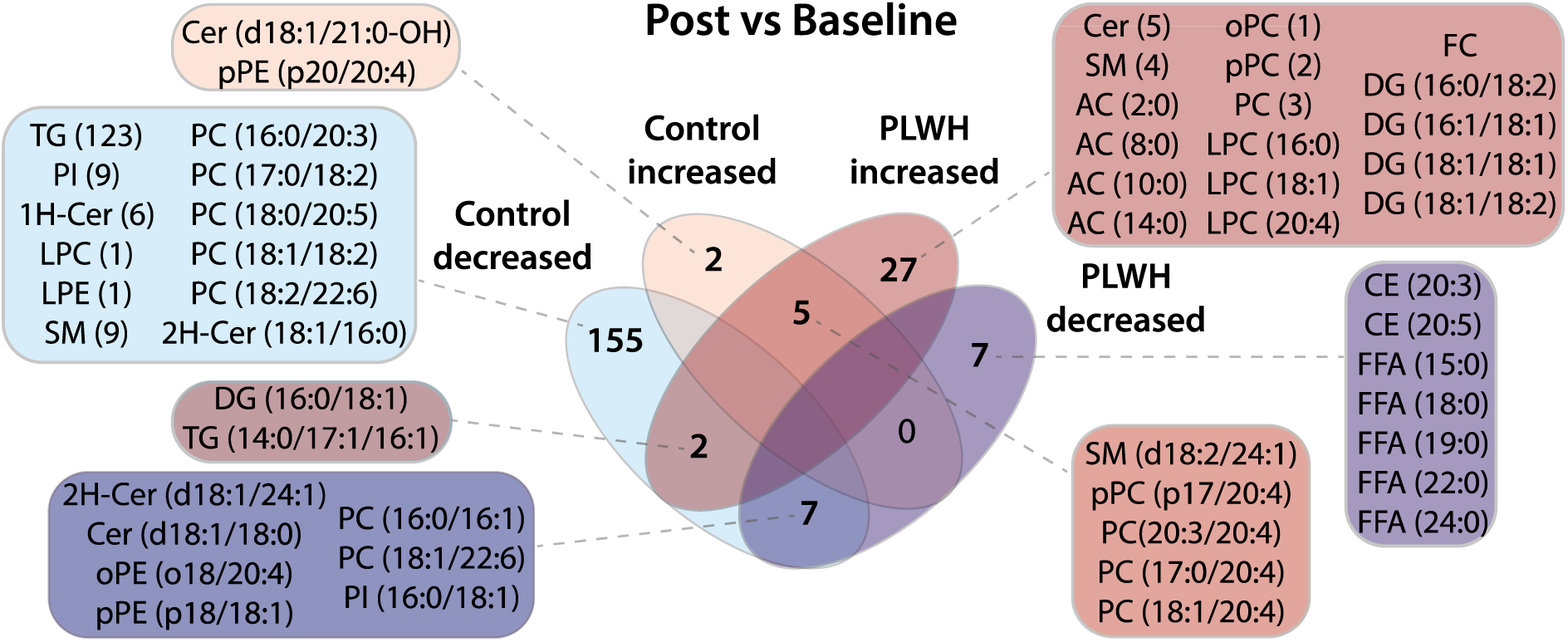
Venn diagram of the comparisons post exercise versus baseline in control subjects and PLWH (as shown in Figure 3). Please see the main text for abbreviations.

Furthermore, we sought to compare the effects of exercise training on plasma lipidome to anthropometric and biochemical measurements, and for this purpose, the summed concentrations of species within each subclass were calculated (**Figure 4**). This analysis revealed that PLWH exhibited lower concentrations of PC and SM relative to control subjects at baseline (as previously mentioned for several lipid species). Plasma lipidome differences were found in concert with higher levels of adiponectin, insulin, HOMA_IR and Adipo_IR, with the latter three exhibiting >2-fold difference in PLWH as compared to control subjects at baseline (**Figure 4a**). Interestingly, the latter differences were equalized post exercise training. Eight weeks of exercise training had no major effects on body fat %, BMI, HDL_C and AST in PLWH as compared to control subjects (**Figure 4a**; **Table 1**). However, exercise training attenuated the reduction of leptin from 3.6-fold at baseline to 1.8-fold in PLWH relative to control, while a slight increase in HOMA_beta (2.2- to 2.6-fold) was noticed (**Figure 4a**). Regarding plasma lipidome profiles, exercise training led to increased concentrations of DG and TG in PLWH as compared to control subjects as well as reduced pPE and total cholesterol levels estimated with traditional clinical measurements.

**Figure 4.**
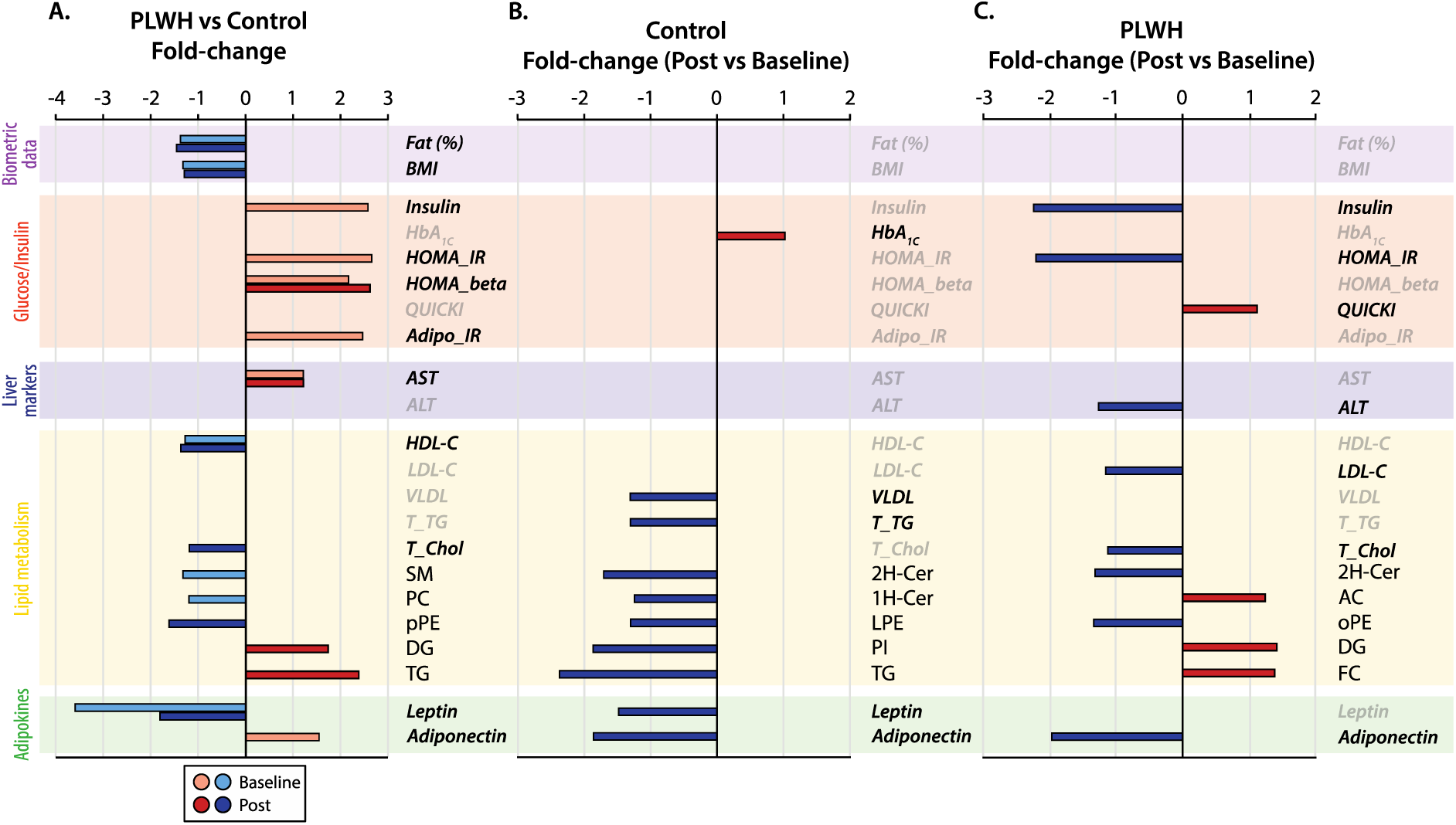
Fold-change variation of significantly altered clinical measurements and lipid subclasses in PLWH versus control subjects at baseline and post exercise (a; unpaired t-test) as well as in paired analysis of individual groups in response to high-intensity training (b,c). Background colors indicate the categories of parameters evaluated (left corner). Please see the main text for the description of lipid subclasses abbreviations.

Clinical and lipidome profiles alterations in response to exercise training by paired analysis revealed a common trend among PLWH and control subjects: decreased concentrations of 2H-Cer and adiponectin, with similar fold-change values, as compared to baseline conditions (**Figure 4b and 4c**). Despite these common alterations, most of the features significantly modulated by high-intensity exercise training were peculiar to each group. In control subjects, decreased concentrations around 2-fold were observed for TG and PI that corresponded to modest reductions of <1.5-fold in LPE and 1H-Cer together with leptin, VLDL and total TG post exercise relative to baseline (**Figure 4b**). Moreover, a small increment in HbA_1c_ was also noticed in control subjects after training. In PLWH, major alterations in post exercise relative to baseline conditions were linked to biochemical parameters, particularly the >2-fold reduction of insulin and HOMA_IR (**Figure 4c**). In addition, modest decreases in ALT, LDL-C, total cholesterol and oPE levels were accompanied by increased DG, FC, AC concentrations and QUICKI in PLWH post exercise.

## Discussion

This study aimed to investigate the impact of high-intensity exercise training for 8 weeks, corresponding to 24 sessions, on the plasma lipidome composition of PLWH as compared to control subjects. In addition to a relatively low number of participants, this non-randomized trial also involved a heterogeneous population regarding clinical conditions presented at baseline, both inter- and intra-groups (**Table SX**). The most significant distinction of PLWH and control subjects at baseline was the contrasting levels of adiponectin, fasting insulin and values of HOMA_IR and Adipo_IR, which were all equalized after 8 weeks of exercise training. Nonetheless, lower levels of leptin, HDL-C, body fat % and BMI combined with elevated AST and HOMA_beta in PLWH relative to control subjects continued from baseline to post exercise. Considering these findings, we will first focus on the major distinctions in plasma lipidome composition between PLWH and control subjects.

One of the most evident features differentiating the plasma lipidome profiles of PLWH and control subjects at baseline were the relatively lower plasma concentrations of several species of PC, pPC and SM, which are all particularly dependent on choline metabolism. Such a negative association of plasma choline-based phospholipids with HIV infection has been reported in earlier lipidomic investigations (Wong et al., 2014; Bowman et al., 2019; Chai et al., 2019; Zhao et al., 2019). Choline is deemed an essential nutrient, and the liver is a major site of choline metabolism as it can be irreversibly oxidized to betaine and plays a central role in the synthesis of hepatic phospholipids (Zeisel and Costa, 2009). The predominant CDP-choline pathway for *de novo* hepatic synthesis (accounting for 70% of liver PC) is largely dependent on choline availability, and PC is the precursor of other choline-containing phospholipids such as the acyl-monoethers oPC and pPC together with LPC and SM (Li and Vance, 2008). In retrospect, liver PC is required for packaging and export of very-low density lipoprotein (VLDL; Nakamura et al., 1967; Buchman et al., 2001; Noga et al., 2002) as demonstrated in both rodent models and human studies showing that choline depletion leads to the development of fatty liver through hepatic TG accumulation (Sherriff et al., 2016). Liver diseases are the number one cause of non-AIDS morbidity and mortality in PLWH (Price and Thio, 2010; Aboud et al., 2007), and its prevalence in HIV-positive population is considerably higher than in general population (Macías et al., 2014; Maurice et al., 2017). Levels of AST were higher in PLWH than control subjects both at baseline and post exercise training, and although not exclusively produced in the liver, AST is measured routinely to evaluate clinical conditions of the liver (Oh et al., 2017). While the main causes of liver diseases in PLWH are still debatable, including the side effects of combined ART (Welzen et al., 2019), there exists recent evidence for significant alterations in gut-liver axis (Martínez-Sanz et al., 2023). For instance, resulting from increased gut microbial choline metabolism, plasma metabolites such as trimethylamine-N-oxide are elevated and associated with atherosclerosis in PLWH (Shan et al., 2018; Srinivasa et al., 2015). Elevated gut microbial choline metabolism leading to choline depletion in PLWH may also trigger hepatic steatosis (Martínez-Sanz et al., 2023), as reported for fatty liver diseases not related to HIV (Spencer et al., 2011; Sherriff et al., 2016). Thus, lower levels of plasma choline-containing phospholipids in PLWH relative to control subjects at baseline may indicate altered gut microbial choline metabolism and/or fatty liver, in both ways suggesting hepatic TG accumulation. Our findings revealed that exercise training in PLWH improved fasting FFA and insulin, HOMA_IR and QUICKI indexes together with a significant decrease in ALT levels, being a more specific marker of the liver’s health status than AST (Oh et al., 2017; Pettersson et al., 2008). Correspondingly, differences in plasma concentrations of choline-containing phospholipids in PLWH relative to control subjects were considerably attenuated after 8 weeks of high-intensity training exercise.

Another intriguing finding was the reduction in several species as well as total concentrations of acyl-monoether PE, particularly pPE or plasmalogen, exclusively after exercise training in PLWH relative to control subjects. The role of circulating plasmalogens in health and disease, particularly in aging, is relatively well studied (Maeba et al., 2015). Thus far, however, the response of circulating plasmalogens to exercise in humans has not been thoroughly investigated. While upon synthesis, plasmalogens are promptly secreted by the liver (Vance, 1990), their reduced plasma concentrations post exercise training may indicate decreased lipoprotein secretion in PLWH relative to control subjects. Alternatively, a significant reduction in LDL-C and total cholesterol was observed in PLWH, but not in control subjects after exercise training, therefore decreased concentrations of pPE could be also linked to reduction in LDL particles.

Our study revealed distinct plasma lipidome alterations between PLWH and control subjects in response to high-intensity exercise training regardless of weight loss. In control subjects, the molecular signatures of exercise training were linked to significant reductions in plasma concentrations of almost all TG alongside PI, 1H- and 2H-Cer. In contrast, PLWH displayed a significant decrease in plasma concentrations of FFA, CE and 2H-Cer together with increased concentrations of DG, AC and FC after exercise training relative to baseline. To a certain extent, these results were consistent with traditional lipid panels revealing reduced concentrations in total TG and VLDL for control subjects and decreased concentrations of LDL-C and total cholesterol for PLWH after exercise training.

Of note, the only biochemical parameter concurrently modulated in both groups after exercise training was the reduced levels of adiponectin. In general, chronic exercise training, particularly involving significant weight loss, leads to opposing responses of the two major adipokines: increased adiponectin combined with decreased leptin (Bouassida et al., 2010; Becic et al., 2018). In the absence of weight loss after exercise, reduced adiponectin levels might reflect improved hepatic clearance rather than secretion by the adipose tissue (Halberg et al., 2009; Zhao et al., 2021). This is the case of lean individuals with non-alcoholic fatty liver disease that display high adiponectin levels relative to controls (Lu et al., 2023), similar to PLWH versus control subjects at baseline, indicating delayed clearance instead of increased adiponectin secretion such as in obesity.

Despite the low number of mutual plasma lipidome alterations after exercise training, an interesting finding was related to the decreased concentrations of seven lipid species, with four of them displaying >10-fold change in both groups. Thus, overlapping reductions in 2H-Cer (d18:1/24:1), PC (16:0/16:1), PC (18:1/22:6) and PI (16:0/18:1) with substantially high fold-change values stand out as molecular signatures of potentially beneficial effects of exercise training in plasma lipidome of PLWH and control subjects. For instance, lactosyl ceramides, such as the 2H-Cer (d18:1/24:1), have been considered markers of inflammation and atherosclerosis (Chatterjee et al., 2021), and in plasma samples their elevated concentrations were associated with both diabetes and CVD risks (Alshehry et al., 2016; Chew et al., 2019).

A concomitant modulation of plasma TG and PI, as observed only in control subjects after exercise training, has also been recently reported by two independent investigations from our group. The latter studies provided evidence for increased plasma TG-PI concentrations during postprandial lipemia elicited by a high-fat meal in healthy women (Yoshinaga et al., 2021) and after 6 weeks of statin washout in subjects with familial hypercholesterolemia (Cerda et al., 2023). Importantly, reduced postprandial lipemia and fasting TG concentrations are consistently reported as hallmarks of both exercise and statin use (Weintraub et al., 1989; Freese et al., 2014; Alvarez-Jimenez et al., 2022). Given the transient episode of hypertriglyceridemia that occurs several times each day (Tremblay et al., 1990; Chapman, 2007), lowering postprandial lipemia by chronic exercise training is likely to be even more relevant than fasting TG concentrations in the context of cardiovascular diseases. In contrast to a lack of changes in body weight, insulin or HOMA_IR from baseline to post exercise, reduced levels of adiponectin and leptin were noticed after 8 weeks of high-intensity exercise training in control subjects. Whether the reduction of two of the most important adipokines is directly or indirectly associated with plasma TG-lowering effects of exercise training is currently unknown. Although supported by our previous data, a direct link between plasma TG and PI concentrations to VLDL and/or postprandial TG-rich lipoproteins levels in response to exercise still needs further mechanistic assessments.

While decreased fasting TG concentrations appears as one of the most often reported effects of chronic exercise training on circulating lipids (Durstine et al., 2002; Trejo-Gutierrez and Fletcher, 2007; Mann et al., 2014), our data showed no apparent evidence for significant TG pool alterations in PLWH despite improved fasting insulin levels, HOMA_IR and QUICKI. A reasonable explanation for the lack of TG pool alterations in PLWH could be related to tissue-specific improvement in insulin resistance. On the one hand, reduced fasting insulin by endurance exercise training in healthy men was demonstrated to significantly increase lipoprotein lipase activity (Bergeron et al., 2001), which would increase VLDL catabolic rates thus lowering plasma TG concentrations. On the other hand, a marked decrease in fasting insulin levels could also result in increased hepatic VLDL secretion (Sparks et al., 2013) and increased production rates as observed after exercise training (Shojaee-Moradie et al., 2016). Thus a delicate balance between the VLDL’s production, secretion and catabolism likely influenced by hepatic and/or peripheral insulin resistance/sensibility status could explain the distinct plasma TG concentration responses to exercise training in PLWH and control subjects.

By far the most remarkable effect of exercise in PLWH was the slight but significant reductions in both total cholesterol and LDL-C, features that were not achieved by control subjects and are seldom apparent in short-term interventions (Wang and Xu, 2017; Durstine et al., 2002). The exact mechanism for such a reduction in LDL-C and total cholesterol levels is not firmly established, but there is evidence for the role of LDL receptor – via reduced levels of proprotein convertase subtilisin/kexin type 9 (PCSK9) prompted by exercise training – for the hepatic clearance of cholesterol-rich particles (Kamani et al., 2015). Consistent with these data, two cholesteryl esters (CE) were found in reduced concentrations, while a slight increase in free cholesterol (FC) was also observed. The latter effects could be attributed to the unusual nature of nonalcoholic fatty liver diseases, where the hepatic synthesis of FC may be uncoupled to CE production (Min et al., 2012). It is worth mentioning that reductions in LDL-C and total cholesterol occurred in concert with lower levels of ALT in PLWH after exercise training.

Although to a lesser extent as compared to LDL-C and total cholesterol, plasma lipidome responses to 8 weeks of exercise training in PLWH could also be perceived as health-promoting alterations. For instance, concentrations of several FFA species were found modestly reduced in PLWH post exercise training. Fasting FFA concentrations are not only elevated in obesity, insulin resistance, non-alcoholic liver diseases and type 2 diabetes mellitus, but also deemed a critical risk factor for cardiovascular disease development (Henderson, 2021). Such reductions in fasting FFA concentrations with chronic exercise training have been reported by several studies (Smart et al., 2016; Thompson et al., 2010; Shojaee-Moradie et al., 2007), although it is currently unclear whether a broad range of exercises (e.g. endurance or resistance training) would invariably lead to similar outcomes (Henderson, 2021). In contrast to FFA, the concentrations of several short- to medium-chain acylcarnitines (AC; 2:0, 8:0, 10:0 and 14:0) increased in PLWH after exercise training. Generally, increased concentrations of AC are associated with insulin resistance and diabetes, reflecting incomplete mitochondrial beta-oxidation and efflux of these compounds into the circulation (Mihalik et al., 2010). However, increased plasma concentrations of AC are also observed in metabolic challenges linked to high lipolytic rates and thus elevated substrate availability for beta-oxidation such as in exercise and starvation (Carlin et al., 1986; Soeters et al., 2009). Earlier studies have provided strong evidence for the release of acetylcarnitine (i.e. AC (2:0)) (Hiatt et al, 1989; Friolet et al., 1994) during high-intensity exercise training, whereas modern metabolomic investigations have linked production and efflux of medium-chain AC to beneficial functions of muscle tissue during moderate intensity exercise training (Lehmann et al., 2010).

Lastly, PLWH after exercise training displayed higher plasma concentrations for all DG species reported in this study. Increased concentrations of DG and ceramides in both skeletal muscle and liver are deemed putative mediators of lipid-induced insulin resistance (Timmers et al., 2008; Petersen and Shulman, 2017). On the other hand, increased DG plasma concentrations are less obvious, but more likely reflect vascular metabolism (e.g. higher lipolytic activity; Lalanne et al., 2003) rather than tissue modifications. Such an increase in plasma DG levels together with decreased TG concentrations were the main findings of a recent targeted lipidomic study evaluating the effects of endurance/resistance exercise in PLWH for 24 weeks (Bowman et al., 2022). Interestingly, the same effect in PLWH was not apparent in their control subjects, which was attributed to differences in lipid metabolism and fatty acid oxidation between populations.

This study has several limitations, including the varying clinical conditions that could not be fully matched with control subjects. From a clinical perspective, PLWH displayed significantly lower BMI, body fat %, leptin and HDL-C levels than control subjects at baseline, while presenting higher levels of adiponectin, AST, fasting insulin and HOMA and Adipo_IR indexes. Although the latter parameters suggest to some degree hepatic inflammation and insulin resistance for PLWH, with improved outcomes after exercise training, insulin sensitivity status was not precisely evaluated (e.g. by hyperinsulinemic-euglycemic clamp). Moreover, even though a few non-specific markers such as AST and ALT were employed in this non-randomized trial, hepatic steatosis was not assessed by accurate imaging methods (e.g. ultrasound or magnetic resonance; Zhou et al., 2019), which ultimately restrained our discussion on liver status to general markers. Despite the high variability in clinical parameters, including inter- and intra-group discrepancies, and possible confounding factors, such as changes in diet composition and medication, the lipidomic data revealed a surprisingly high number of lipid species modulated by exercise training. More importantly, distinct plasma lipidome responses resulted in very specific molecular signatures for each group. The responses of PLWH to high-intensity exercise training have likely revolved around the improvement in insulin sensitivity and hepatic steatosis after 8 weeks, playing a major role in distinguishing plasma lipidome signatures as compared to control subjects. It is uncertain, however, whether an extended program of exercise training for PLWH (i.e. more than 8 weeks) would in fact produce a reduction in VLDL and TG comparable to that detected in control subjects as well as to a 24-week program as reported by Bowman et al. (2022).

In addition to the specific lipidome alterations of each group, this study revealed concomitant modulation in several lipid molecular species suggesting health-promoting effects of short-term high-intensity exercise training. Some of those overlapping plasma lipid species included increased concentrations of PC (17:0/20:4), PC (18:1/20:4), PC (20:3/20:4) and pPC (p17/20:4), all of them linked to a chain of arachidonic acid, together with SM (d18:2/24:1). In contrast, commonly modulated plasma lipid species displaying reduced concentrations included the previously mentioned 2H-Cer (d18:1/24:1), PC (16:0/16:1), PC (18:1/22:6) and PI (16:0/18:1) displaying high fold-change values as well as pPE (p18/18:1), oPE (o18/20:4) and Cer (d18:1/18:0). Collectively, these commonly modulated lipid species represent interesting targets for future lipidomic-based studies evaluating not only the effects of exercise, but perhaps the molecular mechanisms ensuing a healthier plasma lipidome profile. Similarly, a selected number of plasma lipid species was recently reported as potential markers of favorable and deleterious cardiometabolic conditions in the context of aging in both women and men (Carrard et al., 2021). In conclusion, lipidomic analysis examining lipids at the molecular species level may strengthen our ability to understand plasma lipidome variations in health and disease, perhaps leading to an improved patients’ stratification in clinical studies.

## Supporting information

Supplemental_file1_Internal_Standards

Supplemental_file2_Lipid_Data

## Data Availability

All data produced in the present study are available upon reasonable request to the authors.

## Acknowledgments

The authors express their gratitude to the project participants and the laboratory team, with special thanks to the co-authors whose contributions were essential to the successful completion of this project. The authors acknowledge the National Council for Scientific and Technological Development (CNPq) for providing a research scholarship. All individuals mentioned in this section have given their consent to be acknowledged.

## Notes

### Competing Interest Statement

The authors have declared no competing interest.

### Clinical Trial

Brazilian Registry of Clinical Trials (UTN: U1111-1231-1846)

### Funding Statement

This study was funded by the National Council for Scientific and Technological Development (CNPq) and Ministry of Education (MEC)

### Author Declarations

The experimental study procedures were approved by the Institutional Research Ethics Committee under the registration no. 3211855 (Universidade Estadual de Maringa). The experimental trial followed the recommendations outlined by the CONSORT statement for non-pharmacological interventions (Boutron et al., 2008). The clinical trial was registered in the Brazilian Registry of Clinical Trials (UTN: U1111-1231-1846).

## References

1. Aboud M et al. Insulin resistance and HIV infection: a review. Int J Clin Pract. 61(3):463–72. 2007.

2. Alvarez-Jimenez L et al. Effectiveness of statins vs. exercise on reducing postprandial hypertriglyceridemia in dyslipidemic population: A systematic review and network meta-analysis. J Sport Health Sci. 11(5):567–577. 2022.

3. Aristizabal-Henao JJ et al. Nontargeted lipidomics of novel human plasma reference materials: hypertriglyceridemic, diabetic, and African-American. Anal Bioanal Chem. 412(27):7373–7380. 2020.

4. Alshehry ZH et al. Plasma Lipidomic Profiles Improve on Traditional Risk Factors for the Prediction of Cardiovascular Events in Type 2 Diabetes Mellitus. Circulation. 22;134(21):1637–1650. 2016.

5. Becic T, Studenik C, Hoffmann G. Exercise Increases Adiponectin and Reduces Leptin Levels in Prediabetic and Diabetic Individuals: Systematic Review and Meta-Analysis of Randomized Controlled Trials. Med Sci (Basel). 30;6(4):97. 2018.

6. Bergeron J et al. Race differences in the response of postheparin plasma lipoprotein lipase and hepatic lipase activities to endurance exercise training in men: results from the HERITAGE Family Study. Atherosclerosis. 159(2):399–406. 2001.

7. Bouassida A et al. Review on leptin and adiponectin responses and adaptations to acute and chronic exercise. Br J Sports Med. 44(9):620–30. 2010.

8. Buchman AL et al. Choline deficiency causes reversible hepatic abnormalities in patients receiving parenteral nutrition: proof of a human choline requirement: a placebo-controlled trial. JPEN J Parenter Enteral Nutr. 25(5):260–8. 2001.

9. Boutron I et al. Extending the CONSORT statement to randomized trials of nonpharmacologic treatment: Explanation and elaboration. Ann Intern Med.148:295–309. 2008.

10. Bowman ER et at. Altered Lipidome Composition Is Related to Markers of Monocyte and Immune Activation in Antiretroviral Therapy Treated Human Immunodeficiency Virus (HIV) Infection and in Uninfected Persons. Front Immunol. 16:10:785. 2019.

11. Bowman ER et al. Lipidome Alterations with Exercise Among People With and Without HIV: An Exploratory Study. AIDS Res Hum Retroviruses. Jul;38(7):544–551. 2022.

12. Carlin JI et al. Carnitine metabolism during prolonged exercise and recovery in humans. J Appl Physiol (1985). 61(4):1275–8. 1986.

13. Carrard J et al. Metabolic View on Human Healthspan: A Lipidome-Wide Association Study. Metabolites. Apr 30;11(5):287. 2021.

14. Ceccarelli G et al. Physical Activity and HIV: Effects on Fitness Status, Metabolism, Inflammation and Immune-Activation. AIDS Behav. 24(4):1042–1050. 2020.

15. Cerda A et al. Lipidomic analysis identified potential predictive biomarkers of statin response in subjects with Familial hypercholesterolemia. Chem Phys Lipids.257:105348. 2023.

16. Chai JG et al. Association of Lipidomic Profiles With Progression of Carotid Artery Atherosclerosis in HIV Infection. JAMA Cardiol. 1;4(12):1239–1249. 2019.

17. Chapman MJ. Metabolic syndrome and type 2 diabetes: lipid and physiological consequences. Diab Vasc Dis Res. 4 Suppl 3:S5–8. 2007

18. Chatterjee S, Balram A, Li W. Convergence: Lactosylceramide-Centric Signaling Pathways Induce Inflammation, Oxidative Stress, and Other Phenotypic Outcomes. Int J Mol Sci. 12;22(4):1816. 2021.

19. Chaves-Filho AB et al. Alterations in lipid metabolism of spinal cord linked to amyotrophic lateral sclerosis. Sci Rep. 12;9(1):11642. 2019.

20. Chew WS et al. Large-scale lipidomics identifies associations between plasma sphingolipids and T2DM incidence. JCI Insight. 4;5(13):e126925. 2019

21. Chong J et al. MetaboAnalyst 4.0: towards more transparent and integrative metabolomics analysis. Nucleic Acids Res. 46, W486–W494. 2018.

22. Crum-Cianflone N et al. Nonalcoholic Fatty Liver Disease (NAFLD) among HIV-Infected Persons. J Acquir Immune Defic Syndr. 15; 50(5): 464–473. 2009.

23. Diaz SO et al. Exploratory analysis of large-scale lipidome in large cohorts: are we any closer of finding lipid-based markers suitable for CVD risk stratification and management? Anal Chim Acta. 15:1142:189–200. 2021.

24. Durstine JL, et al. Lipids, lipoproteins, and exercise. J Cardiopulm Rehabil. 22(6):385–98. 2002.

25. Edelman EJ et al. The Next Therapeutic Challenge in HIV: Polypharmacy. Drugs Aging. 30(8): 613–628. 2013.

26. Ekroos K et al. Lipid-based biomarkers for CVD, COPD, and aging - A translational perspective. Prog Lipid Res. 78:101030. 2020.

27. Feinstein MJ et al. Patterns of Cardiovascular Mortality for HIV-Infected Adults in the United States: 1999 to 2013. Am J Cardiol. 15;117(2):214–20. 2016.

28. Feinstein MJ, et al. Characteristics, Prevention, and Management of Cardiovascular Disease in People Living With HIV: A Scientific Statement from the American Heart Association. Circulation. 9;140(2): e98–e124. 2019.

29. Finn AV et al. Concept of vulnerable/unstable plaque. Arterioscler Thromb Vasc Biol. 30(7):1282–92. 2010.

30. Freese EC, Gist NH, Cureton KJ. Effect of prior exercise on postprandial lipemia: an updated quantitative review. J Appl Physiol (1985). 1;116(1):67–75. 2014.

31. Friolet R, Hoppeler H, Krähenbühl S. Relationship between the coenzyme A and the carnitine pools in human skeletal muscle at rest and after exhaustive exercise under normoxic and acutely hypoxic conditions. J Clin Invest. 94(4):1490–5. 1994.

32. Gao L et al. Dual mass spectrometry as a tool to improve annotation and quantification in targeted plasma lipidomics. Metabolomics.17;16(5):53. 2020.

33. Gastaldelli A et al. Relationship between hepatic/visceral fat and hepatic insulin resistance in nondiabetic and type 2 diabetic subjects. Gastroenterology, 137(3), 843–854. 2009.

34. Giguère P et al. Getting to the Heart of the Matter: A Review of Drug Interactions Between HIV Antiretrovirals and Cardiology Medications. Can J Cardiol. 35(3):326–340. 2019.

35. Grinspoon A, Carr A. Cardiovascular risk and body-fat abnormalities in HIV-infected adults. N Engl J Med. 6;352(1):48–62. 2005.

36. Han X. Lipidomics for studying metabolism. Nat Rev Endocrinol. 12(11):668–679. 2016.

37. Halberg N et al. Systemic fate of the adipocyte-derived factor adiponectin. Diabetes. 58(9):1961–70. 2009.

38. Henderson GC. Plasma Free Fatty Acid Concentration as a Modifiable Risk Factor for Metabolic Disease. Nutrients. 28;13(8):2590. 2021.

39. Hiatt WR et al. Carnitine and acylcarnitine metabolism during exercise in humans. Dependence on skeletal muscle metabolic state. J Clin Invest. 84(4):1167–73. 1989.

40. Hilvo M et al. Ceramides and Ceramide Scores: Clinical Applications for Cardiometabolic Risk Stratification. Front Endocrinol (Lausanne). 29:11:570628. 2020.

41. Jaggers JR, Hand GA. Health Benefits of Exercise for People Living With HIV. Am J Lifestyle Med. 10(3): 184–192. 2016.

42. Jones R et al. Exercise to Prevent Accelerated Vascular Aging in People Living With HIV. Circ Res. 24;134(11):1607–1635. 2024.

43. Kamani CH et al. Stairs instead of elevators at the workplace decreases PCSK9 levels in a healthy population. Eur J Clin Invest. 45(10):1017–24. 2015.

44. Katz A et al. Insulin sensitivity as assessed by the quantitative insulin sensitivity check index (QUICKI) is effective in identifying insulin-resistant subjects. Diabetes Care, 23(3), 491–497. 2000.

45. Koethe J et al. HIV and antiretroviral therapy-related fat alterations. Nat Rev Dis Primers. 18;6(1):48. 2020.

46. Krishnan A et al. Risk of adverse cardiovascular outcomes among people with HIV and nonalcoholic fatty liver disease. AIDS. 1;37(8):1209–1216. 2023.

47. Kumar S, Samaras K. The Impact of Weight Gain During HIV Treatment on Risk of Pre-diabetes, Diabetes Mellitus, Cardiovascular Disease, and Mortality. Front Endocrinol. (Lausanne). 27:9:705. 2018.

48. Laaksonen R et al. Plasma ceramides predict cardiovascular death in patients with stable coronary artery disease and acute coronary syndromes beyond LDL-cholesterol. Eur Heart J. 1;37(25):1967–76. 2016.

49. Lalanne F et al. Distribution of diacylglycerols among plasma lipoproteins in control subjects and in patients with non-insulin-dependent diabetes. Eur J Clin Invest. 29(2):139–44. 2003.

50. Lehmann R et al. Medium chain acylcarnitines dominate the metabolite pattern in humans under moderate intensity exercise and support lipid oxidation. PLoS One. 12;5(7):e11519. 2010.

51. Li Z, Vance DE. Thematic Review Series: Glycerolipids. Phosphatidylcholine and choline homeostasis. JLR. 49. 1187–1194. 2008.

52. Lu CW et al. Adiponectin-leptin ratio for the early detection of lean non-alcoholic fatty liver disease independent of insulin resistance. Ann Med. 55(1):634–642. 2023

53. Macías J et al. Prevalence and factors associated with liver steatosis as measured by transient elastography with controlled attenuation parameter in HIV-infected patients. AIDS. 1;28(9):1279–87. 2014.

54. Maeba R et al. Plasma/Serum Plasmalogens: Methods of Analysis and Clinical Significance. Adv Clin Chem. 2015:70:31–94. 2015.

55. Mann S, Beedie C, Jimenez A. Differential effects of aerobic exercise, resistance training and combined exercise modalities on cholesterol and the lipid profile: review, synthesis and recommendations. Sports Med. 44(2):211–21. 2014.

56. Martínez-Sanz J et al. A gut microbiome signature for HIV and metabolic dysfunction-associated steatotic liver disease. Front Immunol. 14:14:1297378. 2023.

57. Matthews DR et al. Homeostasis model assessment: insulin resistance and beta-cell function from fasting plasma glucose and insulin concentrations in man. Diabetologia. 28(7), 412–419. 1985.

58. Matyash V et al. Lipid extraction by methyl-tert-butyl ether for high-throughput lipidomics. J. Lipid Res. 49, 1137–1146. 2008.

59. Maurice JB et al. Prevalence and risk factors of nonalcoholic fatty liver disease in HIV-monoinfection. AIDS. 17;31(11):1621–1632. 2017.

60. Mihalik SJ et al. Increased levels of plasma acylcarnitines in obesity and type 2 diabetes and identification of a marker of glucolipotoxicity. Obesity (Silver Spring). 18(9):1695–700. 2010.

61. Min HK et al. Increased hepatic synthesis and dysregulation of cholesterol metabolism is associated with the severity of nonalcoholic fatty liver disease. Cell Metab. 2;15(5):665–74. 2012.

62. Nakamura T et al. Hepatic function tests in heavy drinkers among workmen. Tohoku J Exp Med. 93(3):219–26. 1967.

63. Noga AA, Zhao Y, Vance DE. An unexpected requirement for phosphatidylethanolamine N-methyltransferase in the secretion of very low density lipoproteins. J Biol Chem. 1;277(44):42358–65. 2002.

64. Palella FJ et al. Mortality in the highly active antiretroviral therapy era: changing causes of death and disease in the HIV outpatient study. J Acquir Immune Defic Syndr. 43(1):27–34. 2006.

65. Pettersson J et al. Muscular exercise can cause highly pathological liver function tests in healthy men. Br J Clin Pharmacol. 65(2):253–9. 2008.

66. O’Brien KK et al. Effectiveness of aerobic exercise for adults living with HIV: systematic review and meta-analysis using the Cochrane Collaboration protocol. BMC Infect Dis. 26:16:182. 2016.

67. O’Donnell VB et al. Lipidomics: Current state of the art in a fast moving field. Wiley Interdiscip Rev Syst Biol Med. 12(1):e1466. 2020.

68. Oh RC et al. Mildly Elevated Liver Transaminase Levels: Causes and Evaluation. Am Fam Physician. 1;96(11):709–715.2017.

69. Oliveira VHF et al. Effects of a Combined Exercise Training Program on Health Indicators and Quality of Life of People Living with HIV: A Randomized Clinical Trial. AIDS Behav. 24(5):1531–1541. 2020.

70. Ozemek C, Erlandson KM, Jankowski CM. Physical activity and exercise to improve cardiovascular health for adults living with HIV. Prog Cardiovasc Dis. 63(2):178–183. 2020.

71. Petersen MC, Shulman GI. Roles of Diacylglycerols and Ceramides in Hepatic Insulin Resistance. Trends Pharmacol Sci. 38(7):649–665. 2017.

72. Price JC, Thio CL. Liver disease in the HIV-infected individual. Clin Gastroenterol Hepatol. 8(12):1002–12. 2010.

73. Quiles NN, Piao L, Ortiz A. The effects of exercise on lipid profile and blood glucose levels in people living with HIV: A systematic review of randomized controlled trials. AIDS Care. 32(7):882–889. 2020.

74. Shah, et al. Global Burden of Atherosclerotic Cardiovascular Disease in People Living With HIV: Systematic Review and Meta-Analysis. Circulation. 11;138(11):1100–1112. 2018.

75. Shan Z et al. Gut Microbial-Related Choline Metabolite Trimethylamine-N-Oxide Is Associated With Progression of Carotid Artery Atherosclerosis in HIV Infection. J Infect Dis. 22;218(9):1474–1479. 2018.

76. Sherriff JL, et al. Choline, Its Potential Role in Nonalcoholic Fatty Liver Disease, and the Case for Human and Bacterial Genes. Adv Nutr.15;7(1):5–13. 2016.

77. Shojaee-Moradie F et al. Exercise training reduces fatty acid availability and improves the insulin sensitivity of glucose metabolism. Diabetologia. 50(2):404–13. 2007.

78. Shojaee-Moradie F et al. Exercise Training Reduces Liver Fat and Increases Rates of VLDL Clearance But Not VLDL Production in NAFLD. J Clin Endocrinol Metab. 101(11):4219–4228. 2016.

79. Smart NA et al. Effect of exercise training on liver function in adults who are overweight or exhibit fatty liver disease: a systematic review and meta-analysis. Br J Sports Med. 52(13):834–843. 2016.

80. Sparks, et al. Insulin-dependent apolipoprotein B degradation is mediated by autophagy and involves class I and class III phosphatidylinositide 3-kinases. Biochem Biophys Res Commun. 14;435(4):616–20. 2013.

81. Spencer MD et al. Association between composition of the human gastrointestinal microbiome and development of fatty liver with choline deficiency. Gastroenterology 140(3):976–86. 2011.

82. Soeters MR et al. Muscle acylcarnitines during short-term fasting in lean healthy men. Clin Sci (Lond). 116(7):585–92. 2009.

83. Srinivasa S et al. Plaque burden in HIV-infected patients is associated with serum intestinal microbiota-generated trimethylamine. AIDS. 20;29(4):443–52. 2015.

84. Thompson D et al. Time course of changes in inflammatory markers during a 6-mo exercise intervention in sedentary middle-aged men: a randomized-controlled trial. J Appl Physiol (1985). Apr;108(4):769–79. 2010.

85. Timmers S, Schrauwen P, Vogel J. Muscular diacylglycerol metabolism and insulin resistance. Physiol Behav. 23;94(2):242–51. 2008.

86. Trejo-Gutierrez JF, Fletcher G. Impact of exercise on blood lipids and lipoproteins. J Clin Lipidol. 1(3):175–81. 2007.

87. Tremblay A et al. Effect of intensity of physical activity on body fatness and fat distribution. Am J Clin Nutr. 51(2):153–7. 1990.

88. Tsugawa H, Ikeda K, Arita M. The importance of bioinformatics for connecting data-driven lipidomics and biological insights. Biochim Biophys Acta Mol Cell Biol Lipids. 1862(8):762–765. 2017.

89. Vance JE. Lipoproteins secreted by cultured rat hepatocytes contain the antioxidant 1-alk-1-enyl-2-acylglycerophosphoethanolamine. Biochim Biophys Acta. 16;1045(2):128–34. 1990.

90. Yoshinaga MY et al. Postprandial plasma lipidome responses to a high-fat meal among healthy women. J Nutr Biochem. 97:108809. 2021.

91. Wang Y, Xu D. Effects of aerobic exercise on lipids and lipoproteins. Lipids Health Dis. 5;16(1):132. 2017.

92. Weintraub MS et al. Physical exercise conditioning in the absence of weight loss reduces fasting and postprandial triglyceride-rich lipoprotein levels. Circulation. 79(5):1007–14. 1989.

93. Welzen BJV et al. A Review of Non-Alcoholic Fatty Liver Disease in HIV-Infected Patients: The Next Big Thing? Infect Dis Ther. 8(1):33–50. 2019.

94. Wong, et al. Plasma lipidomic profiling of treated HIV-positive individuals and the implications for cardiovascular risk prediction. PLoS One. 14;9(4):e94810. 2014.

95. Zeisel SH, Costa KA. Choline: an essential nutrient for public health. Nutr Rev Actions. 67(11):615–23. 2009.

96. Zhao W et al. Elevated Plasma Ceramides Are Associated With Antiretroviral Therapy Use and Progression of Carotid Artery Atherosclerosis in HIV Infection. Circulation. 23;139(17):2003–2011. 2019.

97. Zhao S, Kusminski CM, Scherer PE. Adiponectin, Leptin and Cardiovascular Disorders. Circ Res.8;128(1):136–149. 2021.

98. Zhou JH et al. Noninvasive evaluation of nonalcoholic fatty liver disease: Current evidence and practice. World J Gastroenterol. 21; 25(11): 1307–1326. 2019.

