## Supplemental_file1_Internal_Standards for "The effect of short-term and high-intensity exercise training in plasma lipidome profiles of people living with and without HIV"

**Supplemental file Internal Standards**

**Lipid subclasses and respective internal standards with their concentrations in 100 µL.**

| Lipid subclass | Internal Standard | in 50 µL (pM) |
|---|---|---|
| AC | AC (10:0) | 2911.2 |
| FFA | FFA (13:0) | 4665.2 |
| DG | DG (14:0/14:0) | 4875.1 |
| TG | TG (14:0/14:0/14:0) | 3456.9 |
| LPC | LPC (17:1) | 984.9 |
| PC, oPC, pPC | PC (17:0/17:0) | 1640.2 |
| PE, oPE, pPE | PE (17:0/17:0) | 1736.0 |
| PI | PG (17:0/17:0)* | 1664.3 |
| 1H-Cer | 1H-Cer (d18:1/17:0) | 350.1 |
| 2H-Cer | 2H-Cer (d18:1/17:0) | 285.3 |
| Sulf-Cer | Sulf-Cer (d18:1/17:0) | 61.6 |
| Cer | Cer (d18:1/17:0) | 905.9 |
| SM | SM (d18:1/17:0) | 348.6 |
| FC | deuterated FC (d7) | 12732.7 |
| CE | CE (17:0) | 7823.4 |
| Q-10, VitE** | Q-6 | 169.2 |

* a phosphatidylglycerol was used as internal standard and a factor of 0.65 relative to PI was calculated in dilution calibrations. ** the concentrations of VitE were normalized relative to coenzyme Q-6, and therefore should be considered non-quantitative.
